# Left-sided Inhibition Deficit and right-sided Hyperexcitability in Treatment Resistant Bipolar Depression: A TMS-EEG study

**DOI:** 10.64898/2026.08.18.26360692

**Authors:** Eva Oostra, Wianne L. Schipper, Elise B.H. Tans, Eline J. Regeer, Ysbrand D. van der Werf, Philip. F. P. van Eijndhoven, Odile A. van den Heuvel, Eric van Exel, Emile d’Angremont

## Abstract

**Objective:** Disruption of the excitation/inhibition balance may contribute to the pathophysiology of bipolar disorder, with post-mortem studies reporting abnormalities in GABA-receptors, interneurons and inhibitory signaling in prefrontal areas. Transcranial magnetic stimulation with electroencephalography (TMS-EEG) enables *in vivo* assessment of cortical excitability/inhibition. This study examined short-latency intracortical inhibition (SICI) after left- and right-dorsolateral prefrontal cortex (DLPFC) stimulation in bipolar depression (BDep, n=10) and healthy controls (HC, n=22).

**Methods:** SICI (paired-pulse TMS) and excitability (single-pulse TMS) were quantified using local- and global-mean-field-power. Associations with lithium use and between-group differences in TMS-evoked potential amplitudes were also explored.

**Results:** For left-DLPFC stimulation, BDep showed weaker SICI than HC (local: 3.7%±12.5 vs 9.0%±20.3, p=0.05; global: 1.5%±11.7 vs 8.8%±20.6, p=0.10), driven by larger ppTMS responses (weaker inhibition). For right-DLPFC stimulation, BDep showed stronger SICI than HC (local: 17.3%±16.1 vs 0.75%±27.1, p=0.36; global: 16.2%±14.6 vs −1.0%±21.8, p=0.03), driven by a larger spTMS response (enlarged excitability). Stronger right-hemispheric global-SICI was most pronounced in BDep patients not using lithium (17.9%±7.0) vs lithium users (3.9%±11.9) and HC (pFDR=0.03).

**Conclusions:** BDep is characterized by reduced cortical inhibition after left-DLPFC stimulation and enlarged cortical excitability after right-DLPFC stimulation; the latter partly normalized by lithium.

**Significance:** To our knowledge, this is the first study to apply TMS-EEG to the bilateral DLPFC in BDep, revealing distinct patterns of hemispheric dysfunction. These findings warrant replication in larger samples, to further elucidate the underlying pathophysiology and inform the mechanisms of action of neuromodulation treatments, such as rTMS.

## Introduction

Bipolar disorder (BD) affects approximately 2.5% of the general population and is characterized by recurring depressive and (hypo)manic episodes.(McGrath et al., 2023) An increasingly recognized notion in literature is that a disrupted excitation/inhibition (E/I) balance may play an important role in mental disorders, including bipolar disorders (Cao et al., 2021, Sohal and Rubenstein, 2019). It has been suggested that the E/I disbalance in bipolar disorder stems from inhibitory dysfunction (Levinson et al., 2007, Tseng et al., 2024). Post-mortem studies of people with bipolar disorder have reported alterations in interneuron subtypes (Fung et al., 2014, Sakai et al., 2008), abnormalities in GABA_A_ and GABA_B_ receptors (Craddock et al., 2010, Fatemi et al., 2017), and reductions in inhibitory functioning, compared to non-psychiatric subjects (Guidotti et al., 2000).

A consistent finding of transcranial magnetic stimulation combined with electromyography (TMS-EMG) studies was a weaker cortical inhibition in people with bipolar disorder, compared to healthy controls (HC), as indexed by a weaker short-interval intracortical inhibition (SICI) measured with the motor evoked potential (MEP) (Ruiz-Veguilla et al., 2016, Taniguchi et al., 2024). This finding is thought to reflect altered GABA_A_-ergic activity (Ziemann et al., 2015) and was independent of (hypo)manic or euthymic state. However, none of the studies involved patients during a bipolar depression (BDep) episode. Additionally, an inherent limitation of TMS-EMG is that it is limited to corticospinal output of the primary motor cortex (M1), and does not directly capture intercortical network dynamics. Combining TMS with EEG (TMS-EEG) overcomes this limitation by direct measurement of the cortical response to stimulation of a region of choice and thereby provides the opportunity to directly investigate cortical E/I properties. Previous studies applying single-pulse TMS (spTMS) to the left dorsolateral prefrontal cortex (DLPFC) in major depression disorder (MDD) showed differences in TMS-evoked potential (TEP) amplitudes and global mean field power (GMFP), compared to HC (Dhami et al., 2020, Voineskos et al., 2019).

To date, four studies have applied TMS-EEG in people with bipolar disorder, targeting left M1 (Andrews et al., 2016), left premotor cortex (Canali et al., 2017, Canali et al., 2015), or left-DLPFC (Farzan et al., 2010). SICI was found to be weaker in BD compared to controls after M1 stimulation (measured with TMS-EMG). Farzan et al. (2010) investigated the left-DLPFC with TMS-EEG, using long-interval intracortical inhibition (LICI), but found no difference between (state-varying) individuals with BD and HC. None of these studies investigated SICI in DLPFC.

A better understanding of the neurophysiological mechanisms underlying the disrupted E/I balance in BDep could lead to targeted therapeutic interventions, e.g., through the identification of biomarkers predictive of response to repetitive TMS (rTMS). Recent meta-analyses showed potential for left and/or right-DLPFC rTMS in BDep, though no consensus was found for the optimal stimulus location and treatment parameters (Hsu et al., 2024, Konstantinou et al., 2022, Wang et al., 2026). Given the lack of consensus on optimal stimulation site and emerging evidence for lateralized prefrontal dysfunction in BD, bilateral assessment of E/I dynamics is warranted. In this study, we aim to characterize cortical inhibition properties using TMS-EEG in individuals with BDep, targeting left and right-DLPFC. We hypothesize to find a weaker cortical inhibition, after left and right-DLPFC stimulation in individuals with BDep compared to HC. In an exploratory manner, we will investigate the effect of medication use, and the duration and severity of the current depressive episode, on cortical E/I.

## Methods

### Study population

33 HC and 10 participants diagnosed with BD were screened for this study, of whom 22 HC and 10 individuals with BDep were eventually included for analysis (see Supplementary Figure 1). All participants were 18 years or older. The HC sample was recruited as part of the larger TIPICCO trial (TMS-induced plasticity improving cognitive control in obsessive-compulsive disorder; https://clinicaltrials.gov/study/NCT03667807), the BDep sample was recruited via the T-BIDE trial (Efficacy of rTMS treatment for patients with treatment resistant Bipolar Depression; https://www.isrctn.com/ISRCTN16011816). Both trials were approved by the Medical Ethics Committee of Amsterdam University Medical Center (METC reference NL77251.029.21 for T-BIDE; 2018.522 for TIPICCO). Participation was voluntary, and written informed consent was obtained from all participants. The study adhered to the principles outlined in the Declaration of Helsinki and was in accordance with the Medical Research Involving Human Subjects Act (in Dutch: WMO).

Inclusion criteria specific to HC participants were: no psychiatric diagnoses according to the Structured Clinical Interview for DSM-5 Disorders and no psychotropic medication at least for the past 12 months. BDep participants followed the inclusion criteria for the T-BIDE study, which were a Hamilton Depression Rating Scale-17 (HDRS-17) score of at least 17 (moderate to severe depression) during screening, experiencing a treatment resistant depression (at least two medications tried and failed to achieve remission), and no current (hypo)manic episode as indicated by a score<11 on the Young Mania Rating Scale. Participants with contra-indication for MRI or TMS were excluded. See Supplementary Materials for a complete overview of the inclusion and exclusion criteria.

### Study design

This study was conducted on three different days. The informed consent procedure and screening questionnaires were performed on day 1. On the second day, the participants underwent cognitive assessments, i.e., the Tower of London task (for TMS target localization; see Supplementary Materials), during the functional MRI scan (Fitzsimmons et al., 2025). On the third day, participants underwent the TMS-EEG session. See Supplementary Materials for details about the cognitive assessments, MRI and TMS-EEG acquisition.

### TMS-EEG acquisition

TMS pulses were delivered using a MagStim BiStim^2^ stimulator using a figure-of-eight D70 alpha-coil (The Magstim Co.Ltd., Whitland, Wales). In the complete experiment, HC participants underwent stimulation of the left-DLPFC, pre-supplementary motor area, M1 and right-DLPFC, the individuals with BDep underwent stimulation of bilateral DLPFC and left M1. In this study, we focus on the left and right-DLPFC stimulation of both groups.

The resting motor threshold (rMT) was determined using EMG from the right first dorsal interosseous muscle. The rMT was defined as the minimum intensity eliciting ≥50µV MEPs in at least 50% of trials. Left-DLPFC stimulation was guided by neuronavigation (Localite TMS Navigator GmbH, Bonn, Germany), using task-based (Tower of London (Fitzsimmons et al., 2025)) fMRI coordinates, while the right-DLPFC was targeted based on the Beam F3/F4 method (Beam et al., 2009). We applied this in both groups for two reasons: (1) the Beam F3/F4 method aligns with the right-sided targeting approach used in T-BIDE trial, and (2) the task used in the fMRI-guided localization of the left-DLPFC was not designed for right hemispheric targeting (van den Heuvel et al., 2003).

Both the left and right-DLPFC received 153 TMS pulses, i.e., 51 single TMS pulses (1ms, at 120% MT, monophasic waveform), i.e., 51 paired pulses (ppTMS) with an inter-stimulus-interval (ISI) of 2ms, and 51 ppTMS with an ISI of 10ms. These three different pulse paradigms were randomized with inter-trial-intervals between 3-5 seconds. The order of stimulated brain areas was also randomized per participant. In this study, we focus on the spTMS and the 2ms ISI ppTMS to describe SICI. EEG was recorded using a 64-channel EasyCap system (international 10-20 layout), with FCz as reference and AFz as ground.

Data were sampled at 10kHz using a Neuvo 64-channel amplifier (Compumedica Neuroscan, Germany), with electrode impedances kept <5 kΩ. The long axis of the coil was placed at a 45° angle from the midline during stimulation, see also Oostra et al. (2026) and Supplementary Figure 2.

### Preprocessing of acquired TMS-EEG data

The preprocessing steps followed the original TESA toolbox steps (as described by Rogasch et al. (2017)) in EEGLAB running in Matlab 2022b. The electrodes for our region of interests (ROIs) included F1, F3, FC1, and FC3 for the left-DLPFC (Avnit et al., 2023, Chung et al., 2017, Hoy et al., 2021) and contralateral F2, F4, FC2 and FC4 for the right-DLPFC. The time windows were determined *a priori* to account for a relatively large heterogeneity in latencies of the different peaks, but not to overlap with windows of neighbouring peaks of the same polarity. They were determined as follows: for P30 [20-40ms], N40 [30-55ms], P60 [45-75ms], N100 [85-145ms] and P180 [160-220ms].

### Outcome measures and statistical analysis

Data were averaged over trials and over electrodes (either within the ROIs or over all electrodes). The following TMS-EEG outcome measures were extracted per subject and brain area: the area under the curve of the local mean field power (LMFP-AUC; using the ROI electrodes, which is a measure of excitability in the ROI), the AUC of the GMFP (using all electrodes, which is a measure of global excitability), and the amplitude of *a priori* determined TEP peaks. Peaks were defined as a datapoint larger or smaller than +/- five data points within the predefined time-window. If multiple peaks were found, the largest was selected. The time windows used for LMFP- and GMFP-AUCs was [20-270ms].

For our main analysis, we investigated the SICI (see Formula 1 (Farzan et al., 2010)) using the local (LMFP) or global (GMFP) AUC after spTMS and ppTMS.

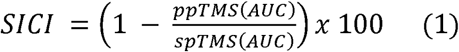

We analysed whether the SICI derived from BDep was weaker compared to HC, for the left- and right-DLPFC locally and globally using the one-sided Wilcoxon’s rank test. Secondly, we investigated the separate ppTMS and spTMS AUCs between BDep and HC. A Spearman correlation test was used to investigate the relation between depression severity, current depression index and bipolar diagnosis index. Depression severity was measured using HDRS-17 with eight additional items covering atypical depression symptoms (Structured Interview Guide for HDRS, with Atypical Depression Supplement; SIGH-ADS (Williams et al., 1988)). Both the SIGH-ADS and the HDRS-17 scores were used for analysis. The Kruskal-Wallis test, with post-hoc Wilcoxon Rank test when *P<*0.05, was used to investigate the influence of medication use on SICI, and separate spTMS and ppTMS AUCs. As an exploratory analysis, we studied the TEP amplitude differences between HC and BDep with the Wilcoxon Rank test. P-values were corrected for multiple testing using FDR correction (only the exploratory analyses, not the primary analyses; since these are hypothesis-driven). For sensitivity analyses, outliers were identified per group for the SICI analysis, using the 1.5*interquartile range (IQR) method and removed.

## Results

### Demographics of the BDep and HC sample

Of the 20 individuals with BDep that were eligible for the TMS-EEG measurements, ten participants declined or were excluded (see Supplementary Figure 1). This left a total of ten participants (mean age 41.1 ± 11.9 years, 60% women). Of the 33 screened HCs, eight participants declined or were excluded, leaving a sample of 25 HCs that completed both the MRI and TMS-EEG visits. The first two datasets were corrupted and therefore unusable; one participant did not complete DLPFC stimulations; the right-DLPFC stimulation was added later in the study. This resulted in a total sample of 22 HC (mean age 36.7 ± 13.6 years, 54.5% women) who received left-DLPFC stimulation, of which eighteen also received right-DLPFC stimulation. The mean rMT of the BDep group (45.4% ± 5.16%) did not differ from rMT of the HC group (42.4% ± 6.44%). All patients used at least two different medications (see Table 1 for the demographics, and Supplementary Table 1 for detailed demographics of the BDep sample). As the BDep sample is small, the variation between medication intake is high and every participant used at least two different classes of psychotropic medication, we decided to focus analyses on medication effects on the associations with lithium usage.

**Table 1.**
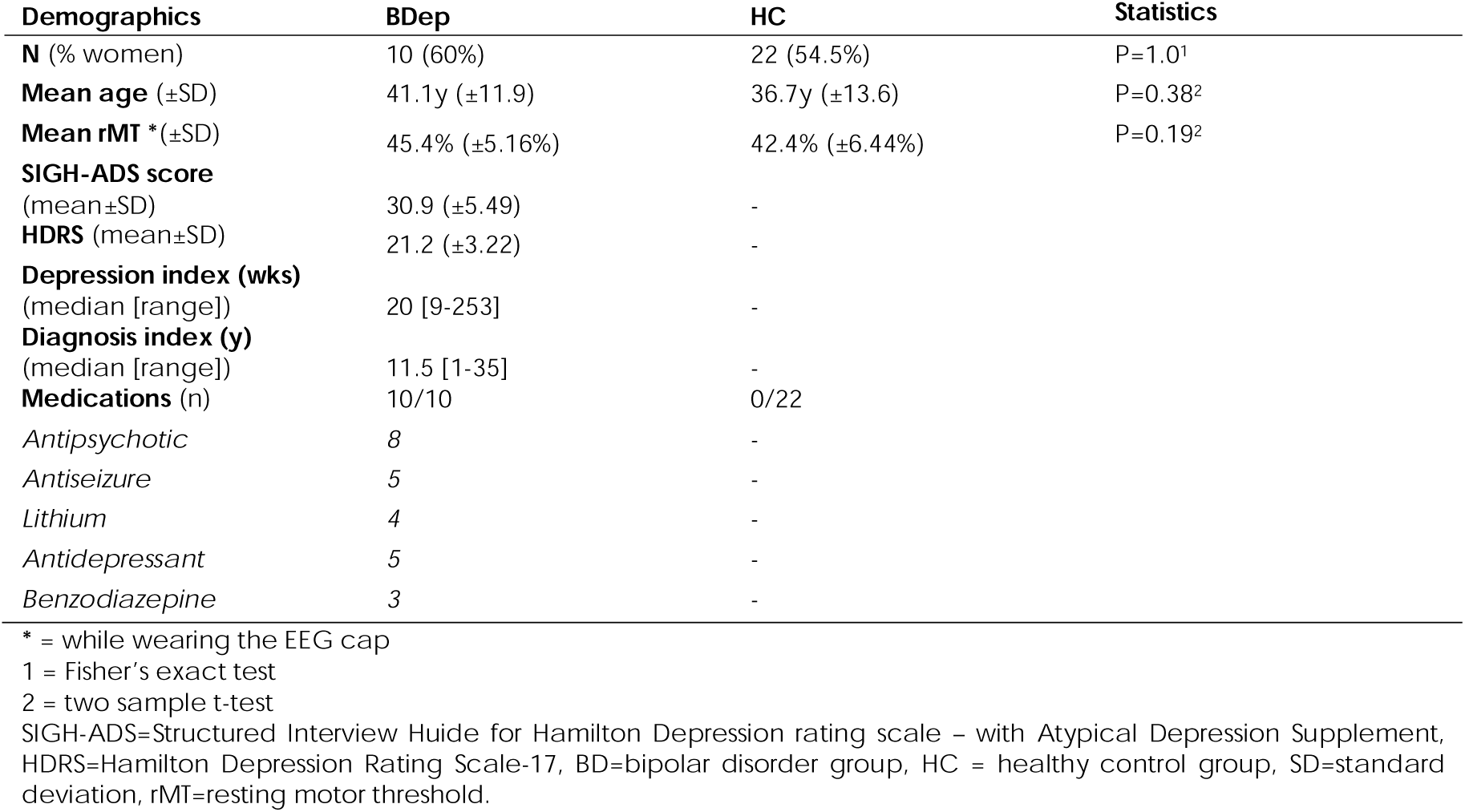
Demographics of study sample.

### Difference in SICI between BDep and HC

After left-DLPFC stimulation, the BDep group showed a near significant weaker local median SICI (3.7% [12.5% IQR]) compared to the HC group (9.0% [20.3% IQR], *W=*150, *P=*0.05), and a weaker global median SICI (1.5% [11.7% IQR]) in comparison to the HC group, but this was not statistically significant (8.8% [20.6% IQR], *W=*142, *P=*0.10). After right-DLPFC stimulation, the BDep group did not show a weaker local or global SICI (17.3% [16.1% IQR] and 16.2% [14.6% IQR], respectively), compared to HC group (0.75% [27.2% IQR], and −1.0% [21.8% IQR], respectively). Additionally, we tested this difference using a *two-sided* Wilcoxon test, and found that the global SICI was statistically significantly stronger in the BDep group (*W=*44, *P=*0.03). See Figure 1 for the visualization of the SICI comparisons, and Table 2 for all the test results.

**Figure 1.**
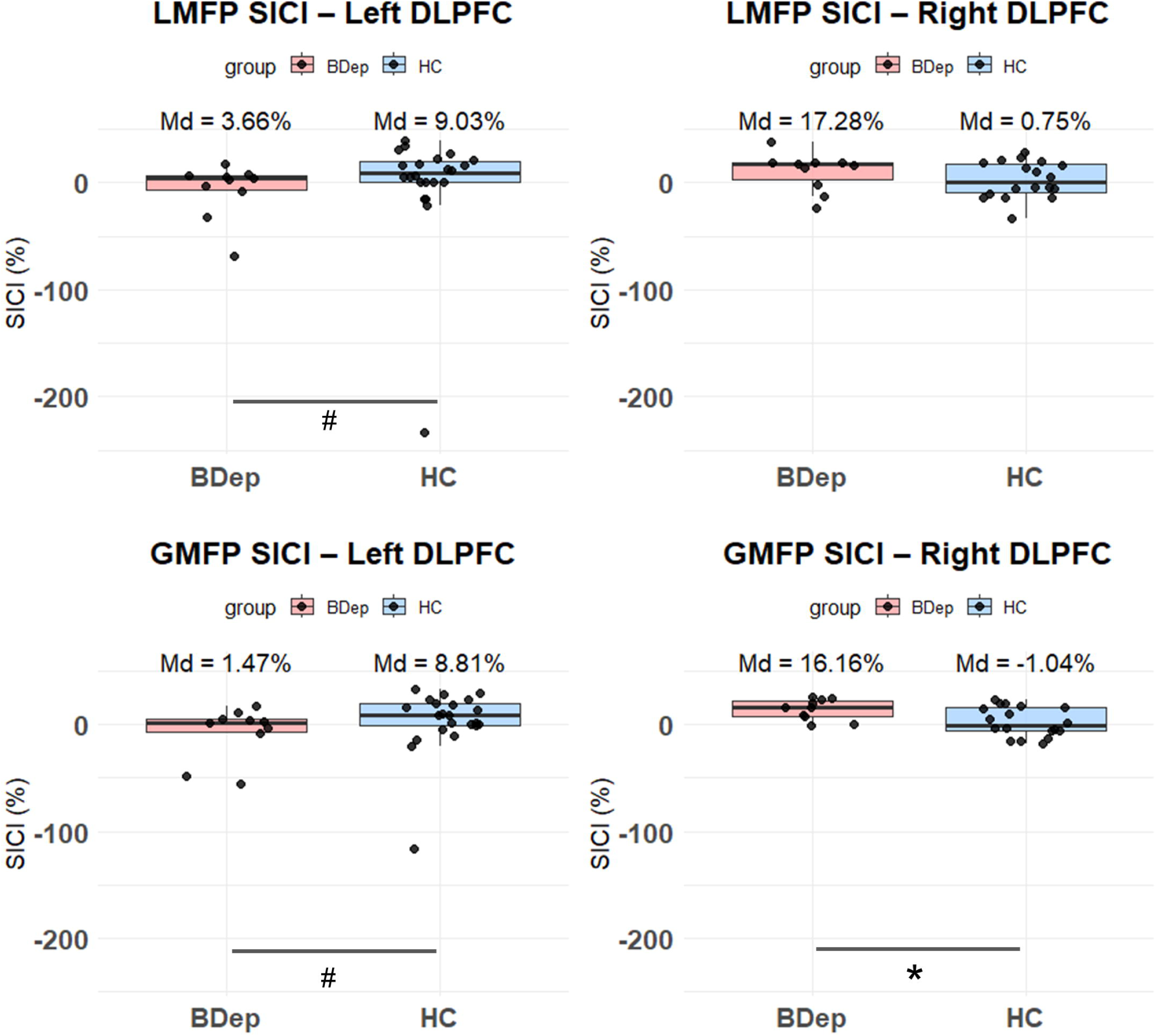
Boxplots of SICI results after left DLPFC and right DLPFC stimulation, on the level of LMFP and GMFP. The top two graphs are SICI derived from LMFP-AUC and the bottom two graphs are derived from GMFP-AUC. The BDep group is visualized in pink on the left of each graph, and HC in blue on the right on each graph. The SICI (ratio) is visualized on the y-axis, with positive values representing facilitation and negative values representing inhibition. Medians are ratio * 100 in percentages for interpretation. # = *P*<0.10, * = *P*<0.05, *SICI (ratio) = 1-(ppTMS_AUC_ / spTMS_AUC_) * 100 with either GMFP-AUC or LMFP-AUC. ppTMS=paired pulse Transcranial Magnetic Stimulation, spTMS=single pulse Transcranial Magnetic Stimulation, AUC=area under the curve, LMFP=local mean field power, GMFP=global mean field power, SICI=short latency intracortical inhibition, DLPFC=dorsolateral prefrontal cortex, BDep=bipolar depression, HC = healthy control*.

**Table 2.**
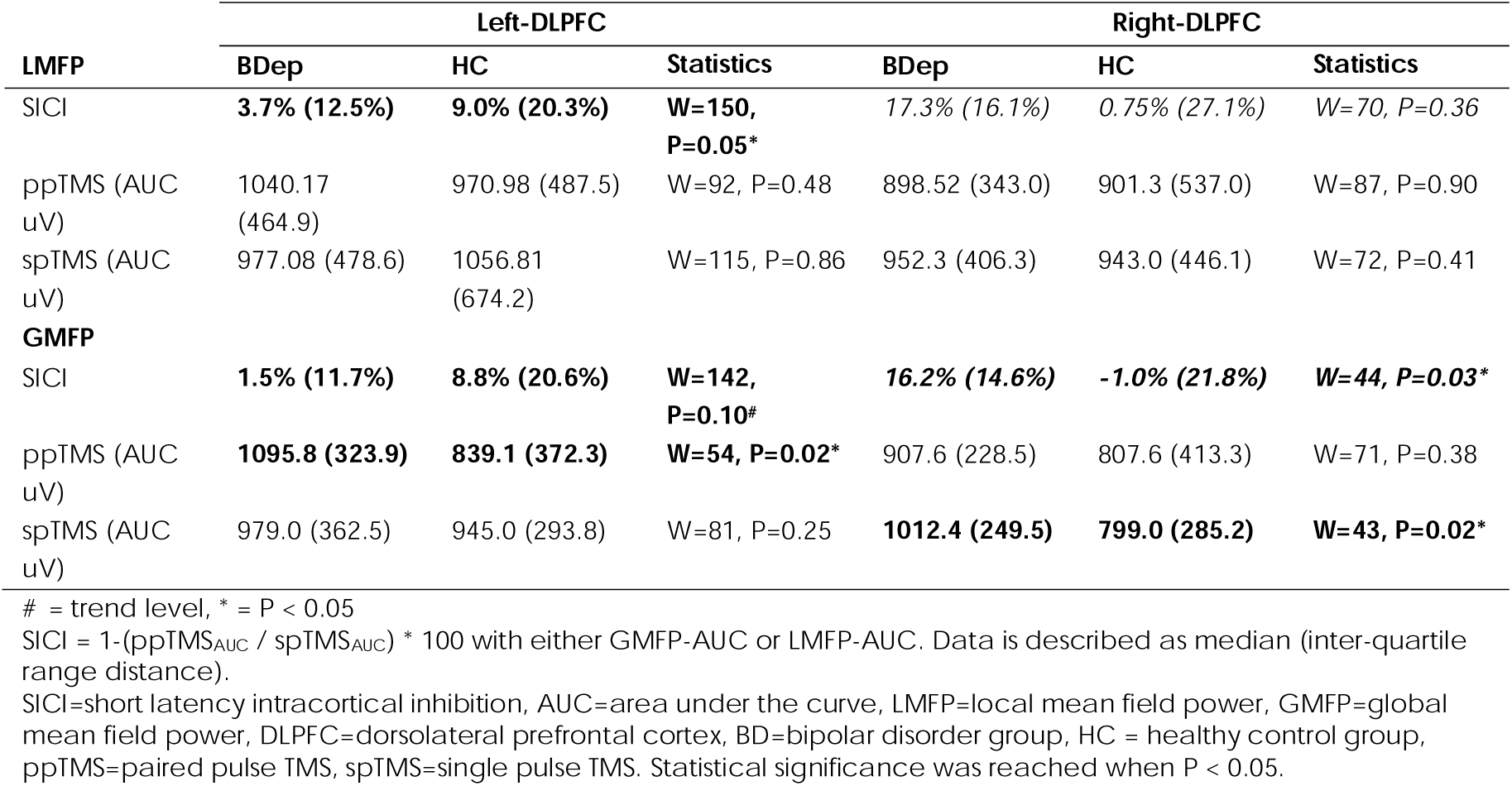
Analysis of the SICI, and LMFP-AUC and GMFP-AUC after left and right DLPFC stimulation. Results are shown in median (IQR), statistics were done using one sided Wilcoxon’s rank test or post hoc using two-sided Wilcoxon’s rank test (in *italic*).

We observed three outliers for the left-sided LMFP and GMFP SICI (BDep, *n=*2 and HC, *n=*1), and one for the right-sided LMFP SICI (BDep, *n=*1). After removing the outliers, the local SICI after left-DLPFC of the BDep was no longer trend-level significantly different (4.7% [5.3% IQR]) compared to HC (10.9% [21.2% IQR], *P=*0.13). Other comparisons were not affected (see Supplementary Figure 3, and Supplementary Materials).

### Difference in AUC of ppTMS and spTMS between BDep and HC

We also investigated if the separate AUCs differed between groups. After ppTMS administration to the left-DLPFC, we found a larger GMFP-AUC in the BDep group (1095.8µV [323.9µV IQR]) compared to HC (839.1µV [372.3µV IQR], *W=*54, *P=*0.02). After spTMS administered to the right-DLPFC, we found a larger GMFP-AUC in the BDep group (1012.4µV [249.5µV IQR]) compared to HC (799.0µV [285.2µV IQR], *W=*43, *P=*0.02). See Table 2 and Supplementary Figure 4 and Supplementary Figure 5. The other comparisons yielded no significant differences.

### Clinical variables

The duration of the current depressive episode, showed a negative, trend-level correlation with the local SICI after left-DLPFC stimulation (*r*=-0.63, *P=*0.05). After one outlier was removed (duration >5 years) the correlation remained negative, but was not trend-level significant anymore (*r*=-0.49, *P=*0.18).

Severity of depression showed a statistically significantly positive correlation with the GMFP-AUC after spTMS administered to the right-DLPFC; for both HDRS score (r=0.85, *P=*0.002) and the SIGH-ADS score (r=0.66, *P=*0.04). Also, the GMFP-AUC after ppTMS applied to the right-DLPFC was positively correlated with HDRS score (r=0.80, *P=*0.005), and with the SIGH-ADS score (r=0.59, *P=*0.08). The right GMFP SICI did not show a statistically significant correlation with HDRS (r=-0.19, *P=*0.60) or with SIGH-ADS scores (r=-0.21, *P=*0.55). Other clinical variables did not show any statistically significance (see Supplementary Table 2).

### Current usage of lithium

When analysing the effect of lithium on SICI, we found a statistically significant effect using the Kruskal-Wallis test (*KW=*6.52, df=2, *P=*0.038) for the global SICI after right-DLPFC stimulation. Pairwise comparisons using Dunn’s post-hoc test indicated that individuals with BDep without lithium (*n=*6) showed a statistically significantly stronger global SICI (17.9% [14.6% IQR]) compared to HC (−1.0% [21.8% IQR], *Z=*2.55, *P=*0.01, pFDR=0.03). Patients who use lithium (*n=*4, 3.9% [11.5% IQR]) showed no significant difference with the HC group (*P=*0.50), see Figure 2 and Table 3. Furthermore, patients who use lithium showed a lower median depression severity score (HDRS: 20.5 [2 IQR], SIGH-ADS: 28.5 [3.5 IQR]), compared to patients without current lithium use (HDRS: 24 [5.25 IQR], SIGH-ADS: 34.5 [7.75 IQR]), although this was not statistically significant (HDRS: *W=*6, *P=*0.24; SIGH-ADS; *W=*5.5, *P=*0.20). When exploring the separate AUCs after ppTMS and spTMS to the right-DLPFC, one can appreciate larger GMFP-AUC after spTMS in the non-lithium users compared to the lithium users and HC. However, this was not statistically significant (Table 3, Supplementary Figure 6, Supplementary Figure 7).

**Figure 2.**
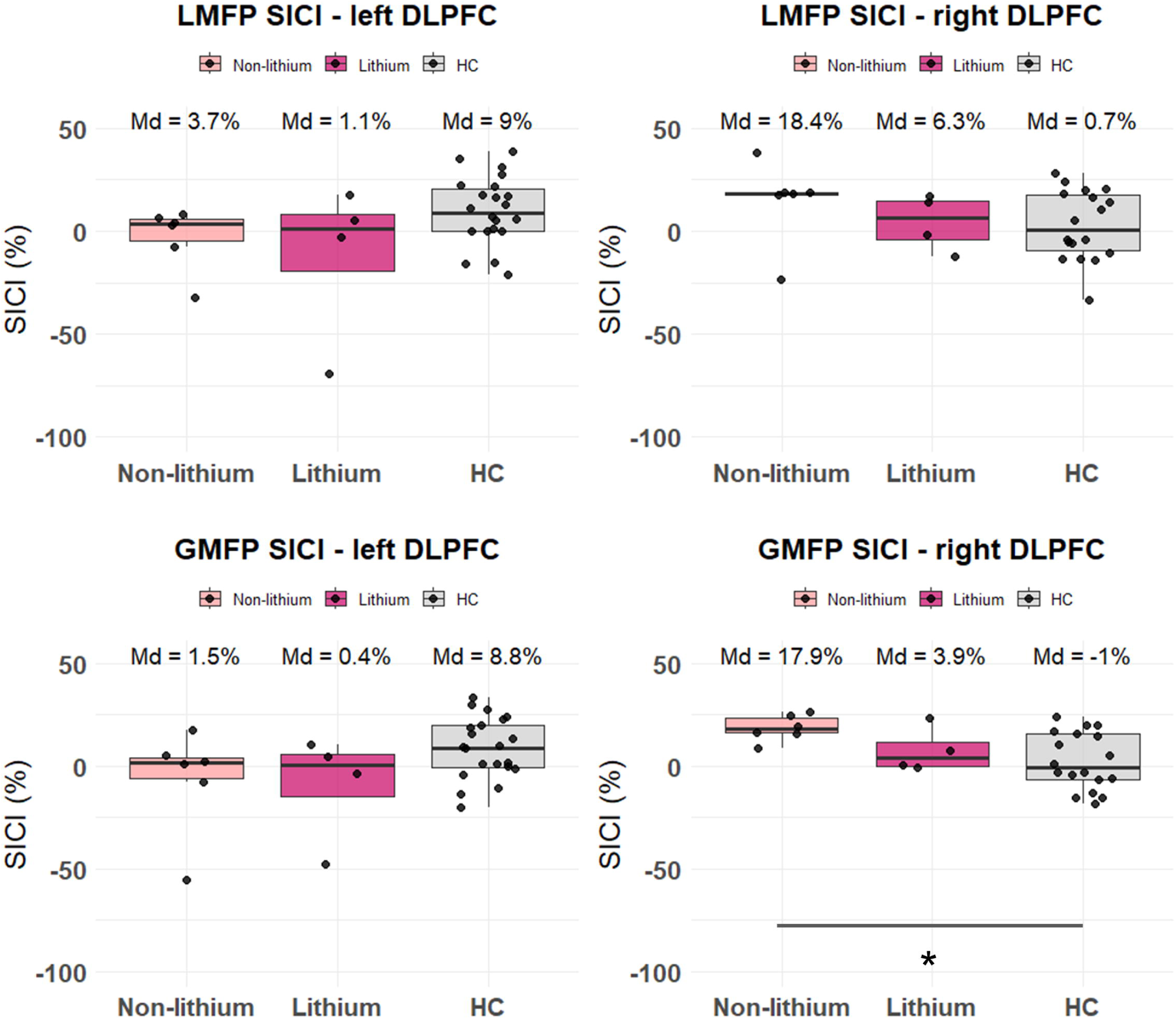
Boxplots of SICI results after left DLPFC and right DLPFC stimulation, with BDep group divided into lithium users and non-lithium users. The top two graphs are SICI derived from LMFP-AUC and the bottom two graphs are derived from GMFP-AUC. The BDep group is visualized on the left of each graph in light pink for non-lithium users and dark pink for lithium users, and HC in gray on the right on each graph. The SICI (%) is visualized on the y-axis, with positive values representing facilitation and negative values representing inhibition. Note: for readability, the y-axis is until −100%. # = *P*<0.10, * = *P*<0.05, *SICI (ratio) = 1-(ppTMS_AUC_ / spTMS_AUC_) with either GMFP-AUC or LMFP-AUC. ppTMS=paired pulse Transcranial Magnetic Stimulation, spTMS=single pulse Transcranial Magnetic Stimulation, AUC=area under the curve, LMFP=local mean field power, GMFP=global mean field power, SICI=short latency intracortical inhibition, DLPFC=dorsolateral prefrontal cortex, BD=bipolar disorder, HC = healthy control*.

**Table 3.**
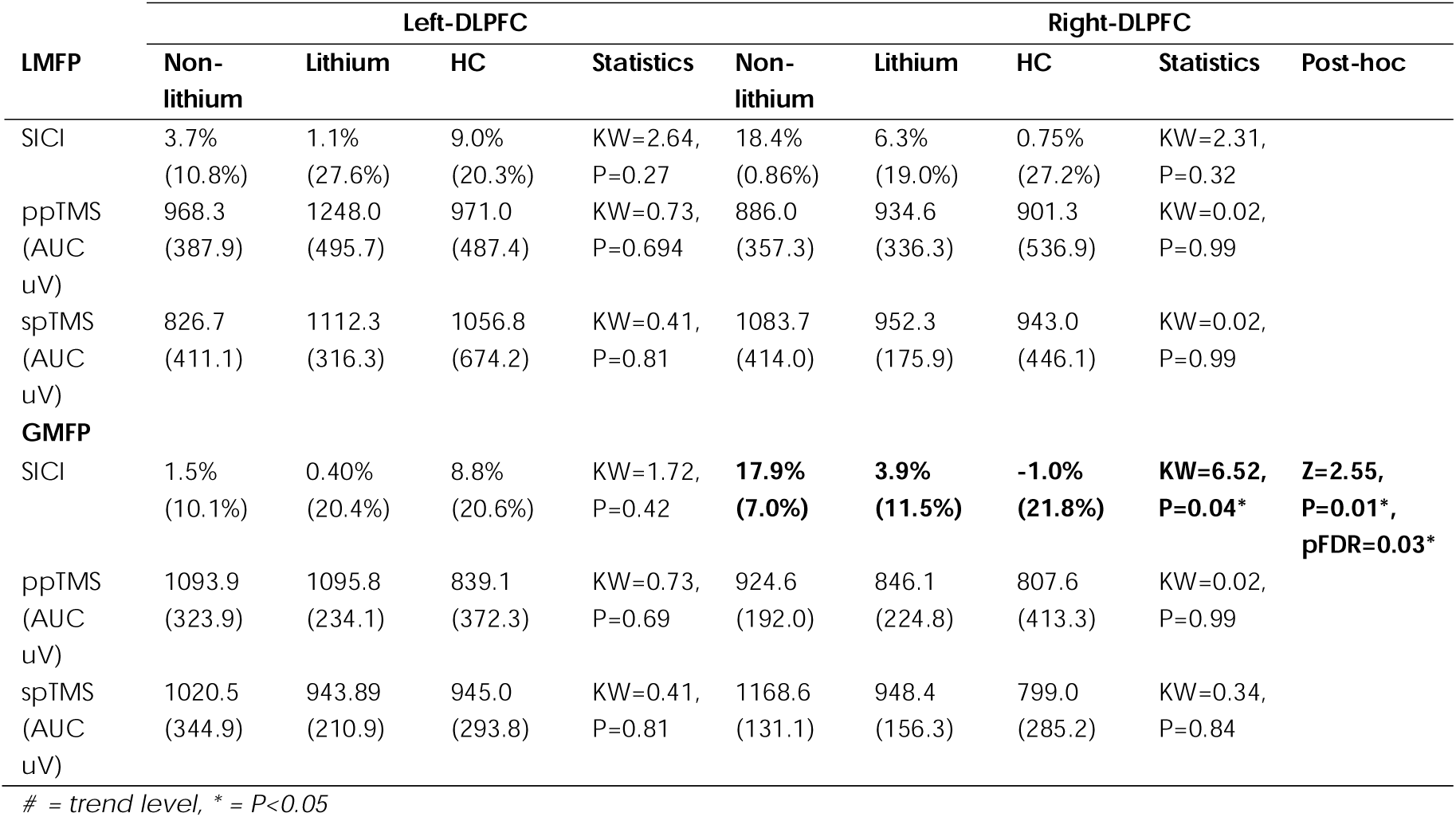
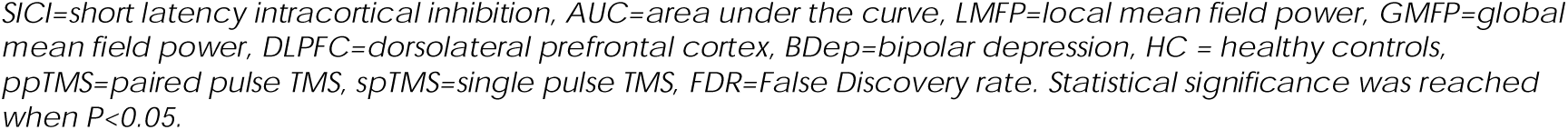
SICI, ppTMS and spTMS AUC values of HC and BDep; BDep divided by lithium usage. Results are shown in median (IQR).

| LMFP | Left-DLPFC |  |  |  | Right-DLPFC |  |  |  | Post-hoc |
| --- | --- | --- | --- | --- | --- | --- | --- | --- | --- |
|  | Non-lithium | Lithium | HC | Statistics | Non-lithium | Lithium | HC | Statistics |  |
| SICl | 3.7%<br>(10.8%) | 1.1%<br>(27.6%) | 9.0%<br>(20.3%) | $KW=2.64$,<br>$P=0.27$ | 18.4%<br>(0.86%) | 6.3%<br>(19.0%) | 0.75%<br>(27.2%) | $KW=2.31$,<br>$P=0.32$ | |
| ppTMS<br>(AUC<br>uV) | 968.3<br>(387.9) | 1248.0<br>(495.7) | 971.0<br>(487.4) | $KW=0.73$,<br>$P=0.694$ | 886.0<br>(357.3) | 934.6<br>(336.3) | 901.3<br>(536.9) | $KW=0.02$,<br>$P=0.99$ | |
| spTMS<br>(AUC<br>uV) | 826.7<br>(411.1) | 1112.3<br>(316.3) | 1056.8<br>(674.2) | $KW=0.41$,<br>$P=0.81$ | 1083.7<br>(414.0) | 952.3<br>(175.9) | 943.0<br>(446.1) | $KW=0.02$,<br>$P=0.99$ | |
| **GMFP** |  |  |  |  |  |  |  |  |  |
| SICl | 1.5%<br>(10.1%) | 0.40%<br>(20.4%) | 8.8%<br>(20.6%) | $KW=1.72$,<br>$P=0.42$ | **17.9%**<br>**(7.0%)** | **3.9%**<br>**(11.5%)** | **-1.0%**<br>**(21.8%)** | **$KW=6.52$,**<br>**$P=0.04^*$** | **$Z=2.55$,**<br>**$P=0.01^*$,**<br>**$pFDR=0.03^*$** |
| ppTMS<br>(AUC<br>uV) | 1093.9<br>(323.9) | 1095.8<br>(234.1) | 839.1<br>(372.3) | $KW=0.73$,<br>$P=0.69$ | 924.6<br>(192.0) | 846.1<br>(224.8) | 807.6<br>(413.3) | $KW=0.02$,<br>$P=0.99$ | |
| spTMS<br>(AUC<br>uV) | 1020.5<br>(344.9) | 943.89<br>(210.9) | 945.0<br>(293.8) | $KW=0.41$,<br>$P=0.81$ | 1168.6<br>(131.1) | 948.4<br>(156.3) | 799.0<br>(285.2) | $KW=0.34$,<br>$P=0.84$ | |

### Difference in response after left and right-DLPFC stimulation

Interestingly, we encountered a lateralization difference in SICI between the groups. In the HC group, we observed a stronger local and global SICI with left-DLPFC stimulation compared to the SICI with right-DLPFC stimulation. The opposite was observed in the BDep group. We analysed this interhemispheric difference by calculating the individual lateralization index (LI) within groups, and compared this between groups (see Supplementary Materials). Locally, we found no SICI lateralization for HC (LI=-0.01), but for individuals with BDep there was a right-sided dominance, which differed on trend-level from HC (LI=-0.12, *W=*193 *P=*0.08). Globally, we found no lateralization for HC (LI=-0.04), but for individuals with BDep there was a right-sided dominance (LI=-0.18, *W=*207, *P=*0.03), which differed statistically significantly from HC (see Table 4).

**Table 4.** Calculation and analyses of the lateralization index (LI). Results are shown in median (IQR).

| LI | BDep | HC | Statistics |
| --- | --- | --- | --- |
| LMFP | -0.12 (0.26) | -0.01 (0.25) | $W=193$, $P=0.08^{\#}$ |
| GMFP | **-0.18 (0.14)** | **-0.04 (0.25)** | **$W=207$, $P=0.03^*$** |

$\#$ = trend level, $*$ = $P<0.05$
$LI = [SICI\text{-}left - SICI\text{-}right]$
SICI=short latency intracortical inhibition, BDep=bipolar depression, HC = healthy control group, LMFP=local mean field power, GMFP=global mean field power. Statistical significance was reached when $P<0.05$ .

### TEP response

As an exploratory analysis, we investigated the difference in TEP amplitude after left and right-DLPFC stimulation between groups. All *a priori* determined TEPs were identified in both groups after left and right stimulation. In the BDep group, we observed a larger N100 (−5.23µV [6.42µV IQR]) after spTMS to right-DLPFC, and a smaller P60 (0.12µV [4.09µV IQR]) after ppTMS to left-DLPFC, compared to the N100 (−3.34µV [1.74µV IQR], *W=*134, *P=*0.037) and P60 (2.09µV [2.28µV IQR], *W=*133, *P=*0.042) of HCs, respectively. These differences were no longer significant after correction for multiple comparisons, see Supplementary Table 4. After left-DLPFC stimulation using spTMS, a positive peak was distinguishable in the BDep group around 85ms, but was absent in HC (see Figure 3, highlighted with an arrow). This peak was absent after right-DLPFC stimulation.

**Figure 3.**
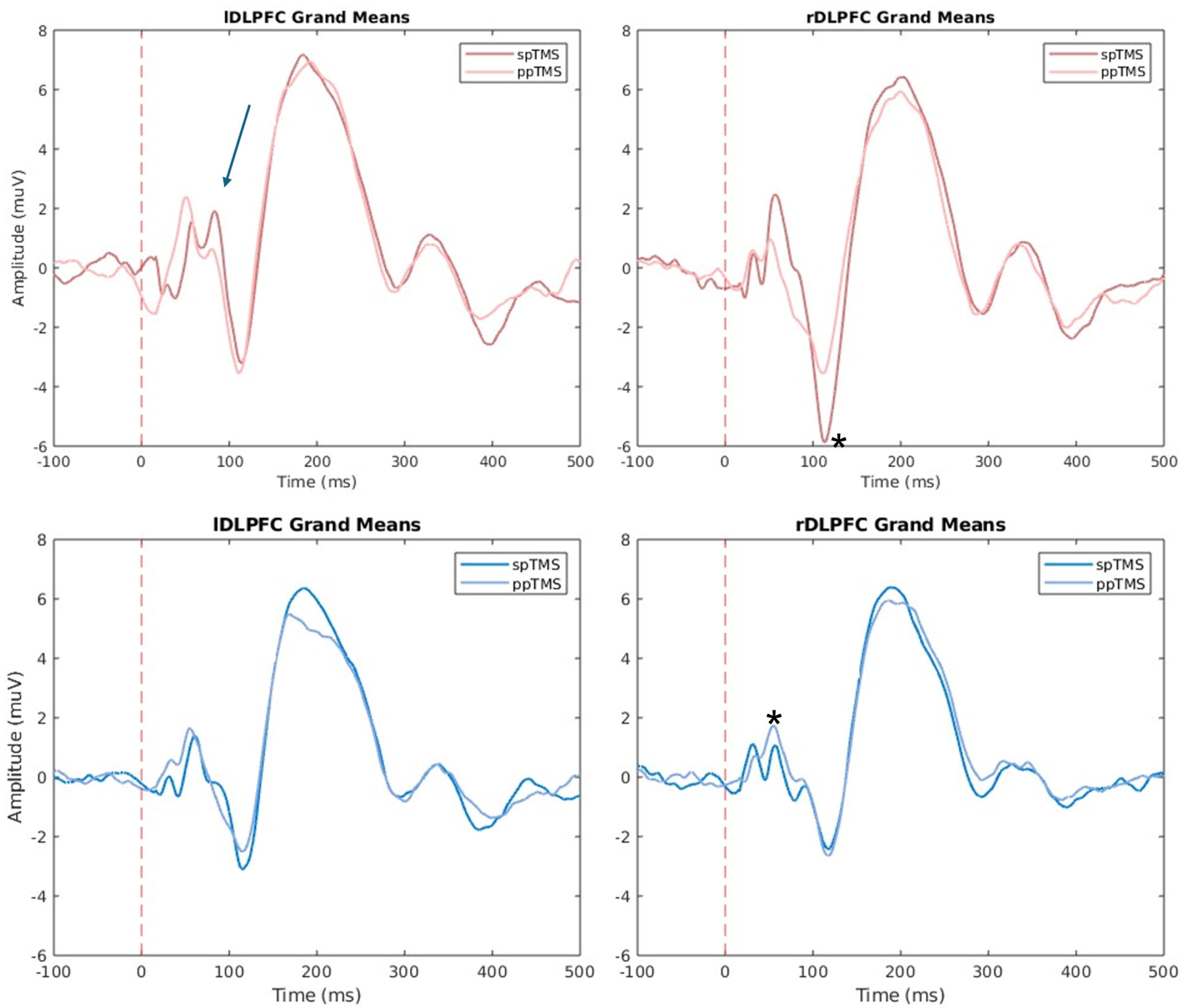
Grand means after spTMS and ppTMS after left and right DLPFC stimulation in BDep and HC. The top two graphs are grand means from the BDep group (pink colors), and the bottom two are grand means from the HC group (blue colors). The dark contrast lines are grand means after spTMS, and the lighter contrast lines are grand means after ppTMS. An arrow is used to identify a TEP that was not determined *a priori*. The asterisk is placed near the peaks that are larger compared to the corresponding peak. *\* = P*≤0.05. *spTMS=paired pulse Transcranial Magnetic Stimulation, DLPFC=dorsolateral prefrontal cortex, BDep=bipolar depression, HC = healthy control*.

## Discussion

In this study we described an investigation of cortical E/I properties in adults with treatment resistant BDep compared to HC, using TMS-EEG. Our findings show a notable lateralization of TMS-EEG responses in BDep. Individuals with BDep (versus controls) showed a trend towards a weaker SICI after left-DLPFC stimulation - confirming our hypothesis -, and a stronger SICI after right-DLPFC stimulation - contradicting our hypothesis. The weaker SICI after left-DLPFC stimulation seemed to be driven by a larger ppTMS response, indicating a weaker cortical inhibition. The stronger SICI after right-DLPFC stimulation is most likely driven by a larger spTMS response, indicating an enlarged excitatory response. The latter appeared to be partially normalized by lithium use.

### Differential cortical response in BDep versus HC

In the left-DLPFC, we found stronger ppTMS response, resulting in a near significance weaker SICI in the BDep group compared to HC – indicating a reduced cortical inhibition. A weaker SICI was also found in studies using TMS-EMG targeting the left M1 in individuals with BD during either a (hypo)manic or euthymic phase (Basavaraju et al., 2019, Basavaraju et al., 2017, Levinson et al., 2007, Ruiz-Veguilla et al., 2016). In MDD, Kinjo et al. (2021) reviewed TMS-EMG results and found that the majority of the studies investigating SICI, showed no differences with HC. Taken together, this suggests that reduced cortical inhibition measured with SICI after left hemispheric stimulation, potentially is a trait-dependent deficit of BD without relatedness to the depressive state (Levinson et al., 2007, Ruiz-Veguilla et al., 2016).

In the right-DLPFC, we found a stronger SICI in the BDep group compared with HC. This was mainly driven by a larger spTMS response in BDep, with no differences in ppTMS responses. This indicates an enlarged excitatory response in BDep. When analysing clinical variables, we found that both spTMS and ppTMS responses correlated with depression severity, while SICI showed no correlation. The association between depression severity and larger spTMS response has been described in patients with MDD as well, but only after left-sided TMS (Cao et al., 2021, Dhami et al., 2020, Voineskos et al., 2019). This mirrored response might point towards a BDep state specific effect. However, the application of right-sided TMS-EEG in MDD is hardly studied and requires further investigation.

Altogether, our findings are in line with a functional cerebral asymmetry model proposed by Moebus et al. (2023). They reviewed fMRI and EEG studies investigating this functional cerebral asymmetry during a (hypo)mania vs. BDep episode, and found prefrontal left-sided dominance (e.g., hyperactivation) during mania and prefrontal right-sided dominance during BDep. Direct neurophysiological measurements, i.e. with TMS-EEG, on a longitudinal scale covering multiple mood states remains to be studied.

### Lithium normalizes enlarged excitation

Our exploratory analysis of medication effects suggested a potential normalizing role for lithium. Patients using lithium demonstrated SICI levels that were more comparable to HC than those not using lithium, after right-DLPFC stimulation. This aligns with previous studies suggesting that lithium may enhance GABAergic inhibitory tone (Malhi et al., 2013). Although our subgroup size (*n=*4) precludes firm conclusions, these findings underscore the importance of accounting for lithium status in future TMS-EEG studies of BD, as it may mask underlying E/I imbalances by stabilizing cortical excitability.

Previous preclinical studies on the effects of lithium on neuronal activity suggested that long-term lithium administration reduces the neuronal response to excitatory agents (Kamp, 2024, Khayachi et al., 2021, Mertens et al., 2015, Santos et al., 2021, Stern et al., 2018). On the macro-level, a systematic review on MRI studies in individuals with BD using lithium, suggests that they show more ‘normalized’ structural brain volumes and functional connectivity in the frontolimbic circuitry, i.e., towards an HC profile (Boere et al., 2026). These regulatory effects of lithium on the cortical response dynamics appears consistent with our findings.

### Individual TEPs in BDep

While exploring the TEP responses of the individual peaks, we found a significantly larger N100 peak after spTMS at the right-DLPFC in the BDep group compared to HC. As the N100 likely depends on GABAergic activity (Premoli et al., 2018) this enlarged amplitude in our BDep sample might reflect a compensatory mechanism for the enlarged excitation measured after spTMS. Another explanation for the enlarged N100-peak could be the influence of lithium usage, as lithium appears to enhance GABA-ergic activity (Malhi et al., 2013).

A surprising finding was the extra peak between P60 and N100 after left-DLPFC spTMS in the BDep group, i.e., a positive peak around 85ms (P85), which was absent in HC. To the best of our knowledge, this peak has not been reported before in TMS-EEG studies. One study investigated sensory gating in individuals with schizophrenia and BD using auditory evoked potentials, with paired auditory clicks with 500ms interval (Patterson et al., 2009). They also reported a P85 peak in the BD group and found that it correlated with perceptual anomalies, which differentiated the BD group from the schizophrenia group (Patterson et al., 2013, Patterson et al., 2009). However, our study did not involve auditory stimuli with 500ms intervals (i.e. 2ms ISI for ppTMS and 3-5 seconds between trials), making it unlikely that the P85 component in our study is derived from the same mechanism described Patterson et al. (2013). Since this work is the first TMS-EEG study reporting this P85 peak in BDep, more research is needed to investigate the underlying mechanisms and its clinical relevance.

### Left vs right

As this is the first study to investigate TMS-EEG targeting both the left and right-DLPFC in BDep, and one of the few that applies TMS-EEG to the right-DLPFC in general, comparison with earlier literature is challenging (Cao et al., 2021, Taniguchi et al., 2024). One example of right-sided TMS-EEG, is the study of Dhami et al. (2020), that directly compared TMS-EEG applied to left and right-DLPFC in youth diagnosed with MDD. They found differences in TEP amplitudes after left (larger P200) and right-DLPFC (larger N100) stimulation compared to controls. They describe a functional asymmetry: the right-DLPFC shows excessive default-mode connectivity, while the left-DLPFC shows excessive cingulate connectivity in MDD (Dhami et al., 2020).

A limitation within our study concerns the difference in localization methods between hemispheres. The left-DLPFC was localized using task-based fMRI, and the right-DLPFC was localized via Beam F3/F4. It is plausible that different parts of the DLPFC were stimulated – e.g. more anterior with Beam F3/F4 compared to fMRI targeting (Kinjo et al., 2024) – resulting in stimulation of different connecting networks. This methodological discrepancy precludes from firm conclusions when comparing left-DLPFC stimulation with right-DLPFC stimulation in our sample. However, the left-right differences appear to be mirrored between BDep and HC, suggesting that the observed asymmetry is not solely attributable to targeting differences. This differential response to TMS on the left and right-DLPFC may reflect lateralized neurophysiological processes. Further investigation is clinically relevant, as it may provide insight into the underlying pathophysiology of BDep, and the mechanisms through which rTMS utilizes its effect when targeting the left or right-DLPFC (Garcia-Toro et al., 2001).

### Strengths, limitations and future recommendations

To our knowledge, this study is the first to investigate both left and right-DLPFC with TMS-EEG in individuals with BD, specifically with a current treatment resistant BDep. As this work represents an exploratory sub-study embedded within a larger clinical trial, our sample size is relatively small – creating a higher risk of Type-2 errors. Results should therefore be interpreted with caution. Even so, several statistically significant effects were observed, indicating that the emerging patterns may be meaningful despite the limited sample.

Another limitation is the heterogeneity in medication use within the BDep group. It is well established that GABAergic medications such as benzodiazepines can influence cortical excitability, measured with spTMS (Minzenberg and Leuchter, 2019, Premoli et al., 2018). Our sample size was too small to statistically control for all medication classes. Nevertheless, this is the first *in vivo* experiment to assess differential neurophysiological effects of lithium usage and non-usage on TMS responses.

This study shed light on the neurophysiological responses after TMS applied to the bilateral DLPFC in individuals with BDep. Our findings showed similarities with findings from earlier research in individuals with current mania, or euthymic state (Taniguchi et al., 2024), and were opposing the findings from TMS-EEG studies in MDD (Kinjo et al., 2021). To disentangle whether our findings are trait- or state-dependent, a larger sample sized study with longitudinal TMS-EEG measurements is warranted. Such a study could visualize neurophysiological changes between individual mood states. This might not only lead to neurophysiological understanding of BD, but also to optimized target locations for treatment options, e.g. rTMS, for all debilitating mood states.

## Conclusion

Altogether, these findings suggest an asymmetrical alteration of the prefrontal cortical response dynamics in BDep. Left-DLPFC stimulation resulted in weaker cortical inhibition, right-DLPFC stimulation in stronger cortical inhibition, as a result of hyperexcitability in response to spTMS. The latter is positively associated with depression severity, and seems to be partially restored by lithium. Replication is needed in a larger BDep sample, to confirm these left/right prefrontal E/I differences and whether these are indicative for neuromodulation treatment options.

## Data availability

The data that support the findings of this study are available from the corresponding author, upon reasonable request.

## Acknowledgements

We thank all the participants for contributing their time and energy to science. Additionally, we thank all the research interns and assistants who’ve supported the study throughout the years, and Chris Vriend for the mental and technical support on the MRI scanning part.

## CRediT Authorship contribution statement

**Eva Oostra:** Conceptualization, Data curation, Formal analysis, Investigation, Methodology, Validation, Project administration, Resources, Visualization, Writing – original draft, Writing – review & editing. **Wianne Schipper:** Methodology, Software, Investigation, Writing – review & editing. **Elise Tans:** Methodology, Software, Investigation, Writing – review & editing. **Eline Regeer:** Writing – review & editing. **Ysbrand D. van der Werf:** Conceptualization, Project administration, Resources, Writing – review & editing, Validation, Supervision. **Philip van Eijndhoven:** Writing – review & editing. **Odile A. van den Heuvel:** Conceptualization, Funding acquisition, Writing – review & editing, Supervision, Validation, Project administration, Resources. **Eric van Exel:** Conceptualization, Funding acquisition, Formal analysis, Writing – review & editing, Supervision, Validation, Project administration, Resources. **Emile d’Angremont:** Conceptualization, Data curation, Formal analysis, Software, Investigation, Methodology, Validation, Project administration, Resources, Visualization, Writing – original draft, Writing – review & editing, Supervision.

## Declaration of generative AI and AI-assisted technologies in the writing process

During the preparation of this work the author(s) used ChatGPT and Microsoft CoPilot in order to improve the readability and language of the manuscript (grammar checks and shortening of sentences). After using this tool/service, the author(s) reviewed and edited the content as needed and take(s) full responsibility for the content of the published article.

