## Supplementary Figure 1 for "Left-sided Inhibition Deficit and right-sided Hyperexcitability in Treatment Resistant Bipolar Depression: A TMS-EEG study"

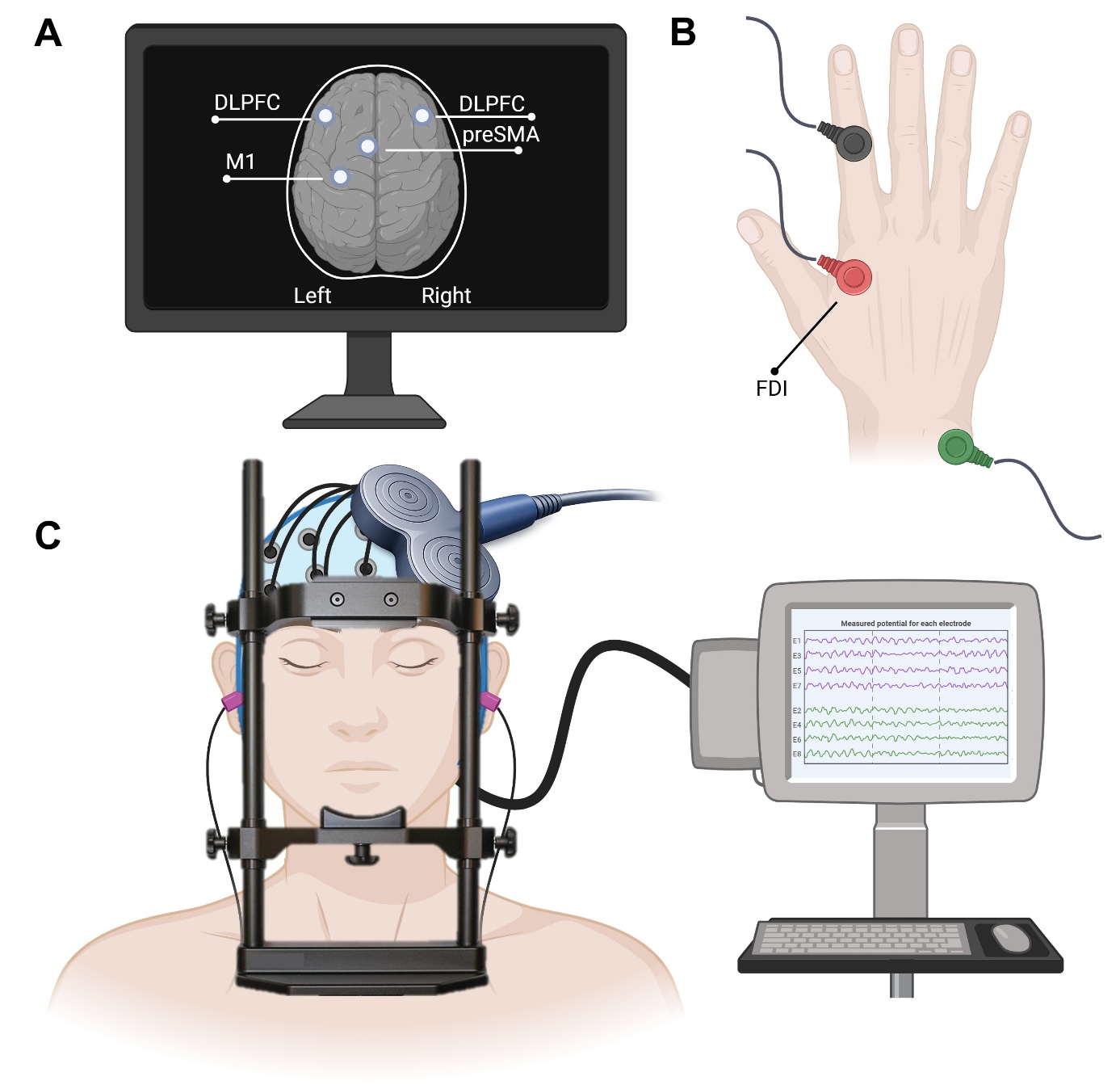

**Figure S1. Experimental set-up of the TMS-EEG session. Adapted from (Oostra et al., 2026).**

A) During the TMS-EEG measurement, neuronavigation is used to localize the brain areas of interest (left M1, preSMA, and bilateral DLPFC). (B) Parallel to M1 stimulation with TMS-EEG, EMG is used to measure the corresponding MEPs. On the right hand, the red electrode is placed on the first dorsal interosseous (FDI), the black electrode is placed on the proximal interphalangeal joint of the index finger, and the green electrode on the pisiform bone. (C) During stimulation, the participant’s head is resting on a chin-rest, with a band around the head to give support. The eyes are closed, and active noise canceling through white noise is played via earbuds. Created in https://BioRender.com.

*TMS=transcranial magnetic stimulation, EEG=electroencephalography, M1=primary motor cortex, preSMA=presupplementary motor area, DLPFC=dorsolateral prefrontal cortex, EMG=electromyography, MEP=motor evoked potential.*
