## Supplementary Figure 2 for "Left-sided Inhibition Deficit and right-sided Hyperexcitability in Treatment Resistant Bipolar Depression: A TMS-EEG study"

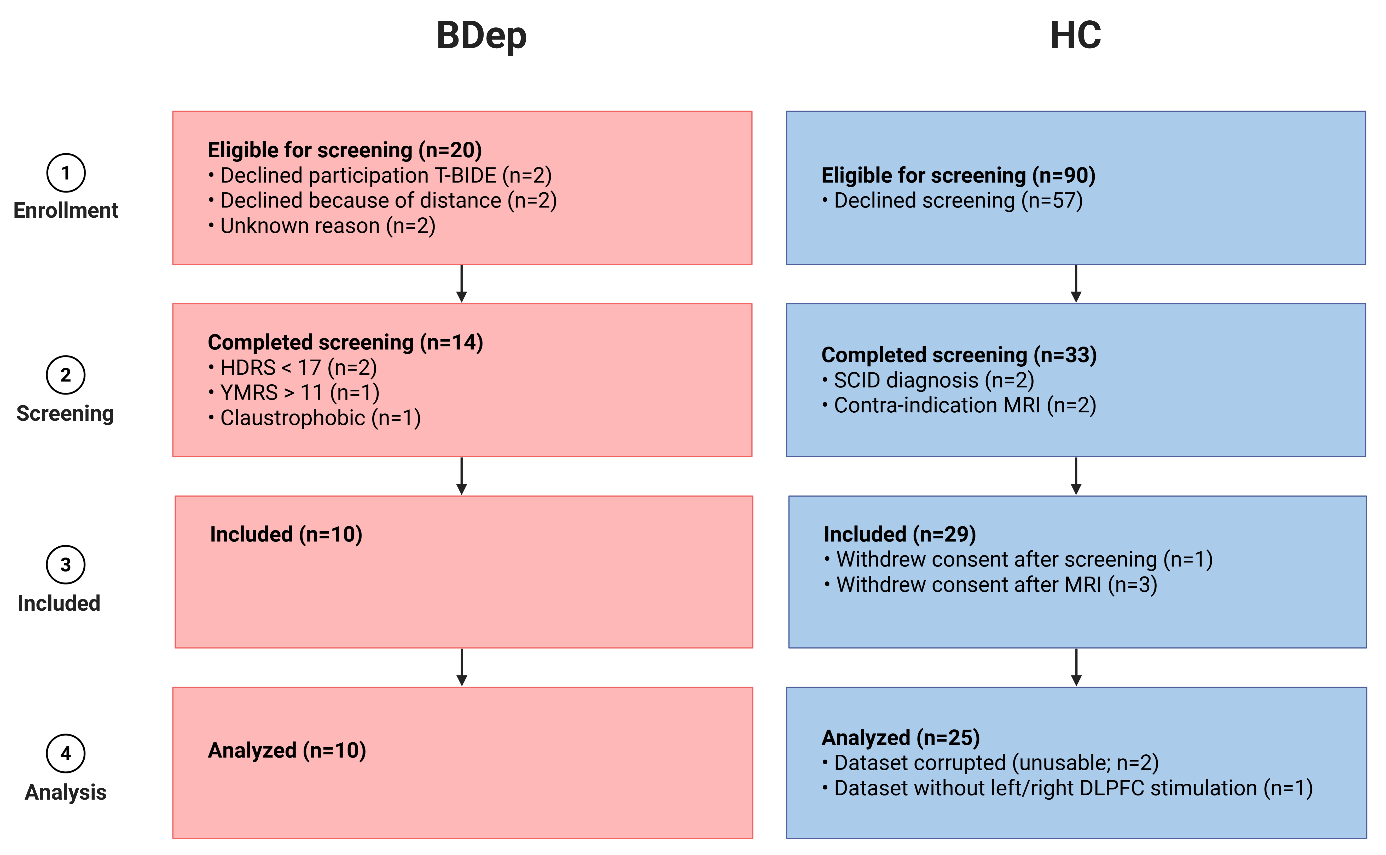

**Figure S2. Flowchart of enrollment and screening of participants.** Created in https://BioRender.com.

*BDep = Bipolar Depression, HC = healthy controls, T-BIDE = clinical trial “Efficacy of rTMS for treatment resistant Bipolar Depression”, TMS-EEG= Transcranial magnetic stimulation with electroencephalography, HDRS = Hamilton depression rating score (17 items), YMRS = Young mania rating scale, SCID = Structured Clinical Interview for DSM-5 Disorders, MRI = magnetic resonance imaging, DLPFC = dorsolateral prefrontal cortex*
