## Supplementary Figure 3 for "Left-sided Inhibition Deficit and right-sided Hyperexcitability in Treatment Resistant Bipolar Depression: A TMS-EEG study"

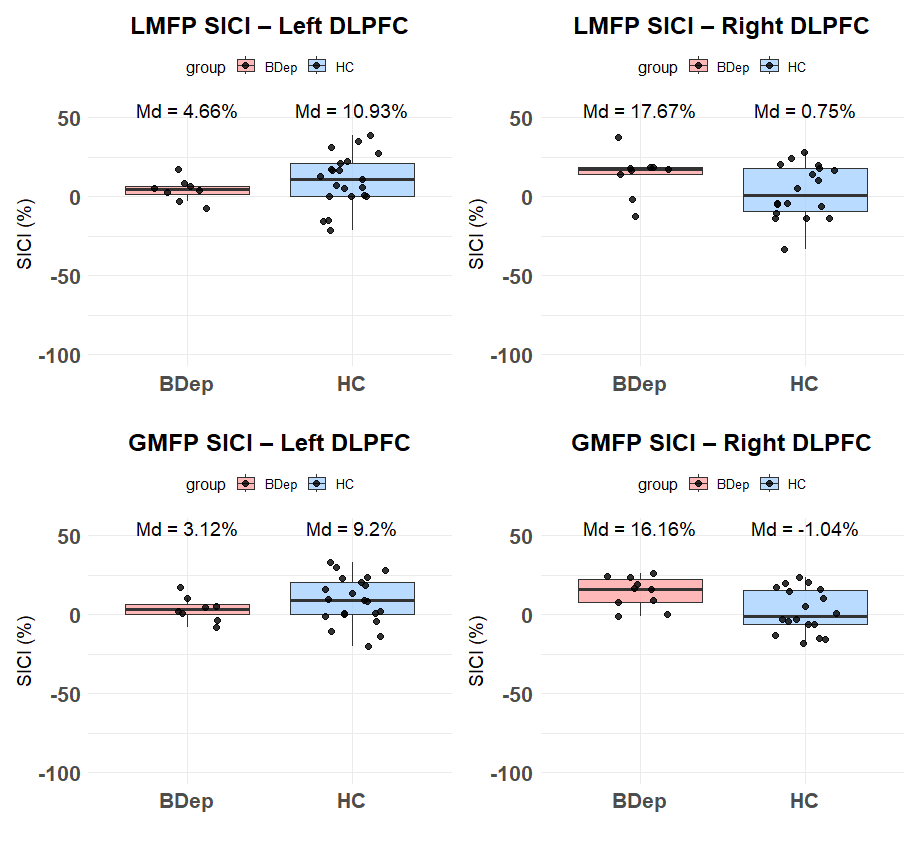

*

**Figure S3. Boxplots of SICI after left and right-DLPFC stimulation, without outliers.** The top two graphs are SICI derived from LMFP and the bottom two graphs are derived from GMFP. The BDep group is visualized in pink on the left of each individual graph, and HC in blue on the right. The SICI (%) is visualized on the y-axis.

*LMFP=local mean field power, GMFP=global mean field power, DLPFC=dorsolateral prefrontal cortex, BDep=bipolar depression, HC = healthy control, Md=median. Statistical significance was reached when p≤0.05.*
