## Supplementary Figure 4 for "Left-sided Inhibition Deficit and right-sided Hyperexcitability in Treatment Resistant Bipolar Depression: A TMS-EEG study"

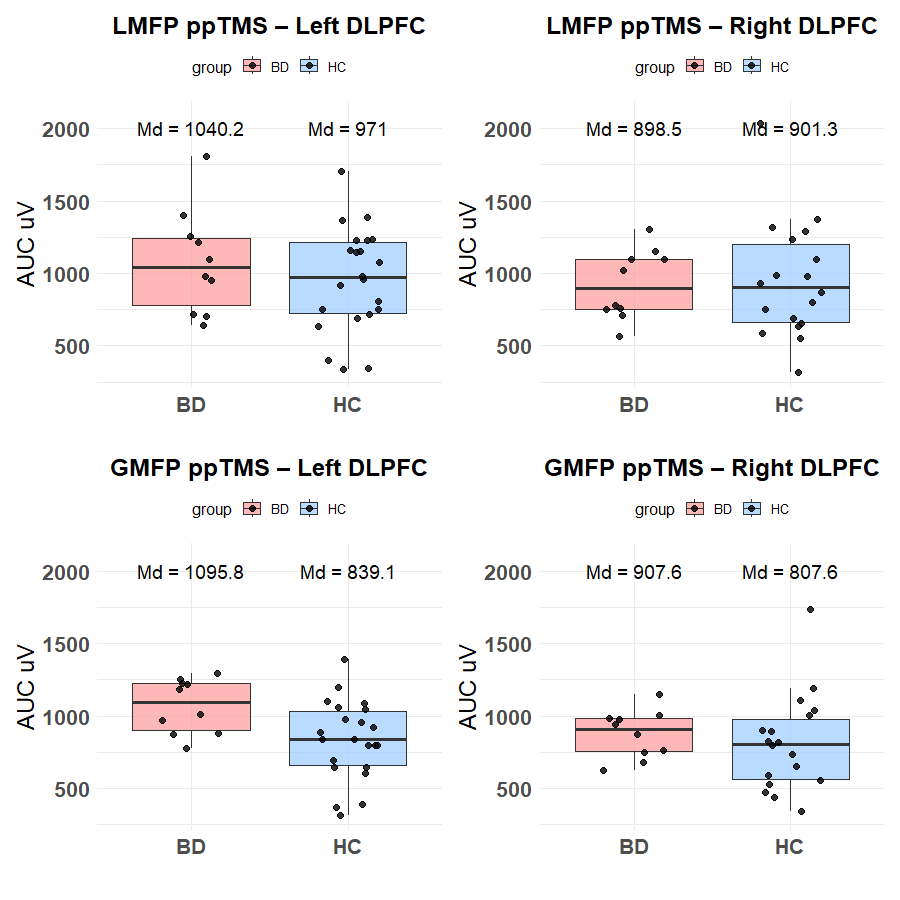

*

**Figure S4. Boxplots of ppTMS after left DLPFC and right DLPFC stimulation, on the level of LMFP and GMFP.** The top two graphs are AUCs derived from LMFP and the bottom two graphs are derived from GMFP. The BD group is visualized in pink on the left of each graph, and HC in blue on the right on each graph. The AUC (µV) is visualized on the y-axis.

*# = trend level, * = p≤0.05.*

*ppTMS=paired pulse Transcranial Magnetic Stimulation, AUC=area under the curve, LMFP=local mean field power, GMFP=global mean field power, DLPFC=dorsolateral prefrontal cortex, BD=bipolar disorder, HC = healthy control, Md=median. Statistical significance was reached when p≤0.05.*
