## Supplementary Figure 6 for "Left-sided Inhibition Deficit and right-sided Hyperexcitability in Treatment Resistant Bipolar Depression: A TMS-EEG study"

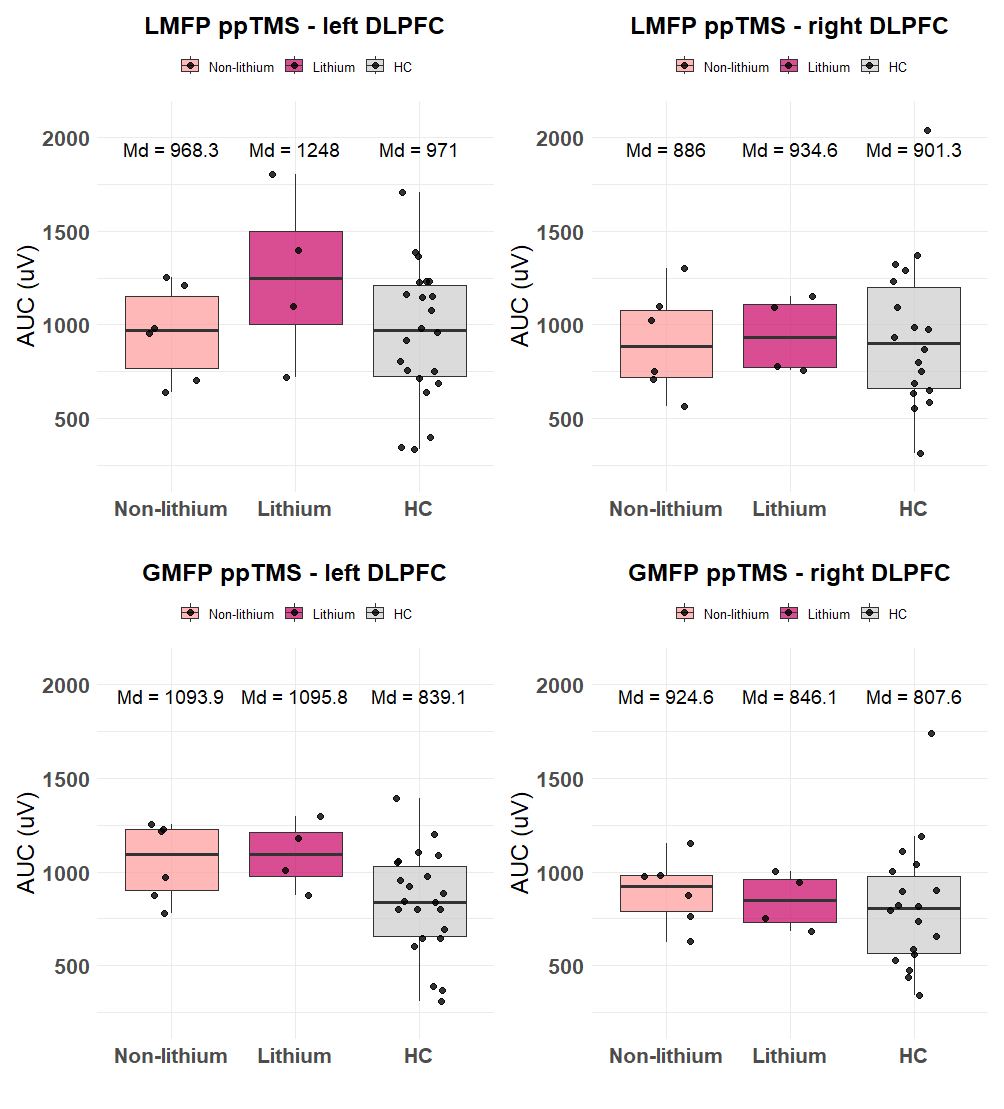

**Figure S6. Boxplots of ppTMS after left DLPFC and right DLPFC stimulation, with BD divided into lithium users and non-lithium users.** The top two graphs are AUCs derived from LMFP and the bottom two graphs are derived from GMFP. The BD group is visualized on the left of each graph in light pink for non-lithium users and dark pink for lithium users, and HC in gray on the right on each graph. The AUC (µV) is visualized on the y-axis.

*# = trend level, * = p≤0.05.*

*ppTMS=paired pulse Transcranial Magnetic Stimulation, AUC=area under the curve, LMFP=local mean field power, GMFP=global mean field power, DLPFC=dorsolateral prefrontal cortex, BD=bipolar disorder, HC = healthy control, Md=median. Statistical significance was reached when p≤0.05.*
