## Supplementary Materials for "Left-sided Inhibition Deficit and right-sided Hyperexcitability in Treatment Resistant Bipolar Depression: A TMS-EEG study"

*A Transcranial Magnetic Stimulation-Electroencephalograhy pilot study*

Eva Oostra* ^1,2,3,4^, Wianne L. Schipper ^1,2,5^, Elise B.H. Tans ^2^, Eline J. Regeer^6^, Ysbrand D. van der Werf^2,5^, Philip. F. P. van Eijndhoven^7,8^, Odile A. van den Heuvel^1,2,5^, Eric van Exel^1,3,4^ & Emile d’Angremont^2,9^

1) Amsterdam UMC, Vrije Universiteit Amsterdam, Dept. Psychiatry, De Boelelaan 1117, Amsterdam, the Netherlands

2) Amsterdam UMC, Vrije Universiteit Amsterdam, Dept. Anatomy & Neuroscience, De Boelelaan 1117, Amsterdam, the Netherlands

3) GGZ inGeest Specialized Mental Health Care, Amsterdam, The Netherlands

4) Amsterdam Neuroscience, Mood, Anxiety, Psychosis, Sleep & Stress program, Amsterdam, The Netherlands

5) Amsterdam Neuroscience, Compulsivity Impulsivity Attention, Amsterdam, the Netherlands

6) Altrecht Mental Health Care Institute Utrecht, and University Utrecht, Department of Clinical Psychology

7) Department of Psychiatry, Radboud University Medical Center, Huispost 961, PO Box 9101, 6500 HB Nijmegen, The Netherlands

8) Donders Institute of Brain Cognition and Behavior, Centre for Neuroscience, PO Box 9104, 6500 HE Nijmegen, The Netherlands

9) Amsterdam Neuroscience, Neurodegeneration program, Amsterdam, the Netherlands

**1.	Methods and Materials**

1.1 *Participants*

*In- and exclusion criteria healthy control sample*

Healthy controls were included when 1) age was between 18-65 years old and 2) no psychiatric, neurodevelopmental or neurological diagnosis was present (assessed using the Structured Clinical Interview for the Diagnostic and Statistical Manual of Mental Disorders IV Axis I disorders (SCID)), or if there was a personal history of DSM-5 diagnosis, except for a diagnosis of depression or anxiety longer than 12 months ago; (3) use of psychotropic medication within the last 12 months; (4) contraindications to MRI such as pregnancy, metal in the teeth, iron in the body and claustrophobia, (5) contraindications to TMS, e.g. epilepsy or family history of epilepsy (first-degree family member), metallic implanted devices, any neurological disorder that causes a lesion in the brain, head trauma resulting in unconsciousness for at least one hour, and previous brain surgery, pregnancy or breastfeeding, syncope, use of pro-convulsive medication or medication affecting the cortical excitability, sleep deprivation, severe heart disease, comorbid substance use/dependence/abuse, resting motor threshold higher than 75% of maximum stimulator output, or no useful motor evoked potential (MEP) eligible.

*In- and exclusion criteria BDep sample*

Patients with BDep were included when 1) age was 18 or above, 2) no other primary diagnosis than bipolar disorder present, 3) HDRS-17 score was >16, 4) no current (hypo)manic episode (YMRS < 12) or at least longer than 12 weeks ago, 5) there was no rapid cycling (> 3 mood swings in the past 12 months), 6) BD type 1 diagnosis accompanied with at least one anti-manic/antipsychotic/mood stabilizer medication, BD type 2 diagnosis *with current antidepressant medication* was accompanied with at least one anti-manic/antipsychotic/mood stabilizer medication, 7) no acute suicidal thoughts, 8) no neurological deficits (tested with the Montreal Cognitive Assessment).

Recruitment of the bipolar sample began after the HC protocol was already halfway completed, which meant that we were bound to the existing HC protocol. This protocol originally applied solely left-DLPFC stimulation, using task-fMRI guided neuronavigation (Oostra et al., 2026, van den Heuvel et al., 2003). When the BDep sample recruitment started, we added right‑hemispheric stimulation to the HC protocol to ensure comparability with the T-BIDE study.

*1.2	Cognitive assessment battery*

The following tasks were conducted before entering the MRI scanner: the Dutch translation of the National Reading Test (to estimate ones IQ), Visual spatial N-back task (to assess working memory), Emotional Stroop task (to measure selective attention and processing speed), and Temporal Discounting and Risk Choice task (to assess risk preferences). After the cognitive assessment battery, participants practiced the computerized Tower of London (TOL) and Stop-signal task (SST) as preparation for the execution during the MRI scan.

*Tower of London task*

The TOL paradigm used in this study was the same as described in Fitzsimmons et al., (Fitzsimmons et al., 2025). The fMRI computerized task consisted of five different planning conditions, varying in difficulty, and one counting condition as the control condition. During the planning conditions the participants were required to plan a sequence of moves to match the beads of the start situation to the goal situation. The five planning conditions differed in the amount of moves needed to reach the goal image; from level 1 (one move needed) till level 5 (five moves needed). During the counting condition, the participant had to count the number of yellow and blue beads presented in the image. The different conditions were presented in a pseudorandomized order, ensuring that each planning condition with level 3 or higher was followed by a counting condition. The stimulus presentation was self-paced, with a maximum response time of 60 seconds; the total task length was fifteen minutes.

*1.3	MRI acquisition*

The MRI scans were done at the Amsterdam UMC, location VU medical center, utilizing a Discovery MR750 3.0T MR scanner, using a 64-channel head coil. The MR images were acquired with 42 ascending slices per volume (slice thickness=3mm, inter-slice gap=0.33 mm, and 3.3x3.3 mm in-plane resolution) and a gradient echo-planar image sequence (TR=2.2 seconds, TS=26 ms, 64x64 matrix, field of view=21.1 cm, flip angle=90º). The head of the participant was immobilized during scanning, to minimize movement artifacts. Participants looked at the screen placed behind the MR scanner, where the tasks were visualized, through a mirror placed on the head coil. Responses of the participants were collected using an MR compatible response box (Current Designs, Philadelphia, PA, USA).

*1.4	TMS-EEG acquisition*

In total, four brain areas were stimulated using one single and two paired pulse TMS paradigms; one paradigm with 2 ms interval (D2; the SICI paradigm) and one with 10 ms interval (D10; the ICF paradigm). In both the D2 and D10 paradigm, the first stimulus was the condition pulse which was set at an intensity of 80% MT, and was followed by the test pulse at 120% MT. The single pulse paradigm contained one test pulse of 120% MT. Each stimulation paradigm consisted of 51 trials, resulting in a total of 153 trials per brain area. The order of the stimulation paradigms was randomized within one brain area (with an inter-trial-interval alternating between 4, 5 and 6 seconds), as well as the order of the brain areas to stimulate per participant. The localization of the brain areas differed; the left DLPFC and left preSMA were localized using the acquired brain areas by the TOL and SST brain activation, respectively. The right DLPFC was localized using the Beam F3/F4 method, and the left M1 localization was done during the MT determination.

*1.5	Preprocessing of (f)MR images*

The task-based fMR images were processed using SPM12 (Wellcome Trust Centre for Neuroimaging London). Functional images were manually reoriented to the structural T1 scan. The first three volumes were discarded and slice timing correction, scanulling (of >2mm of frame to frame displacement), normalization and spatial smoothing (using an 8 mm Gaussian kernel) was carried out during preprocessing. A high-pass filter (128-second cutoff period) was applied to remove low-frequency noise.

For the left DLPFC localization, the local maximum in its area (defined as Brodmann area 9 and 46) during the planning contrast retrieved from the TOL task (all planning conditions > baseline) was defined. If the peak voxel was not on top of a gyrus, the stimulation target was manually placed more on top of the closest gyrus. This was done in the participants T1 scan. For the left preSMA localization, we used the local maximum of the medial gyrus of Brodmann area 6 (anterior to the anterior commissure) during the response inhibition contrast (successful stop trials > successful go trials) of the SST. If no suitable local maximum could be detected, we used the following literature coordinates (in MNI space): Left DLPFC: -40, 28, 30 (7) and for left preSMA: -4, 14, 58 (8). For the right DLPFC, we retrospectively determined the stimulation location on the T1 by extracting the entry and target location from the neuronavigation tool. Once the individualized stimulation coordinate was defined, a 5mm ROI was created, warped from MNI to subject space and overlaid on a T1 MRI scan of the individual participant, allowing navigation to the individualized stimulation location of the left DLPFC and preSMA during the session.

*1.6	EEG preprocessing steps*

TMS-EEG data was pre-processed using a two-step Independent Component Analysis (ICA) procedure in EEGLAB according to the TESA pipeline (Rogasch et al., 2017), running in MatLab 2022b. During the first step of preprocessing, automated removal of bad electrodes was performed in two stages. First, electrodes were removed based on the FlatlineCriterion (SD>5) and the LineNoiseCriterion (SD>4). Second, electrodes were removed based on kurtosis (SD>4). Afterward, epoch segmentation took place (1500ms before and after the TMS pulse), which was followed by a baseline correction using a time frame of 800ms until 110ms before the TMS pulse administration. During the second step of the preprocessing, large TMS pulse and TMS-evoked muscle artifacts were removed by removing 5ms before and 15ms after the TMS pulse administration. This was replaced utilizing cubic interpolation after which the data was down-sampled to 1kHz. Next, automated removal of bad epochs based on joint probability (SD>3) was performed, followed by a visual inspection of the epochs. For details, see Oostra et al., 2026.

**2.	Results**

*2.1	Sensitivity analysis without outliers*

After removing 2 BDep participants from the left DLPFC SICI analysis and 1 HC, we found a local SICI of 4.7% [5.2% IQR] for BDep and 10.9% [21.2% IQR] for HC, that was no longer trend wise significant (W=108, p=0.13). The global SICI of BDep was 3.1% [6.4% IQR] and 9.2% [20.2% IQR] for HC (W=100, p=0.23). After right DLPFC stimulation, only one BDep participant was removed, and solely for the local SICI analysis. The local SICI remained non-significant between BDep (17.7% [4.6% IQR]) and HC (0.8% [27.2% IQR]; W=53, p=0.16). See Figure S7 and S8.

*2.2	Lateralization index (LI)*

To quantify the interhemispheric differences between groups, we calculated the individual lateralization index (LI) per group, for local and global SICI. LI is the difference between left and right DLPFC response (see Formula A). For this, we transformed the SICI to a ratio (SICI/100 = SICIr). When LI≈0 it indicates no lateralization, LI>0 indicates left-sided dominance, and LI<0 right-sided dominance. This was calculated locally (using SICI from LMFP-AUC) and globally (using SICI from GMFP-AUC). The difference LI was tested between groups using a Wilcoxon’s rank test.

$$A) LI={SICIr}_{left}- {SICIr}_{right}$$
