## Supplementary Table 1 for "Left-sided Inhibition Deficit and right-sided Hyperexcitability in Treatment Resistant Bipolar Depression: A TMS-EEG study"

|  |  |  |  |  |  |  |  |  |  |  | **Left DLPFC** | | **Right DLPFC** | |
| --- | --- | --- | --- | --- | --- | --- | --- | --- | --- | --- | --- | --- | --- | --- |
| **Subject** | **BD-type** | **Index depression** | **HDRS** | **SIGH-ADS** | **Lithium** | **Lithium serum (mmol/kg)** | **AD** | **AP** | **AE** | **Benzo** | **SICI LMFP** | **SICI GMFP** | **SICI LMFP** | **SICI GMFP** |
| **01** | BD-2 | 52 | 25 | 38 | n | - | y | n | y | n | -7.63% | 0.95% | -23.77% | 16.43% |
| **02** | BD-2 | 56 | 21 | 28 | y | 0.76 | y | y | n | y | 5.10% | -3.46% | 16.88% | 0.26% |
| **03** | BD-1 | 56 | 18 | 28 | n | - | y | y | n | n | 3.09% | 1.99% | 37.88% | 19.28% |
| **04** | BD-1 | 14 | 24 | 38 | n | - | n | n | y | y | 4.22% | -8.11% | 18.66% | 15.89% |
| **05** | BD-1 | 14 | 24 | 34 | n | - | n | y | y | n | 6.42% | 4.88% | 18.61% | 26.08% |
| **06** | BD-2 | 9 | 21 | 29 | y | 0.33 | n | y | n | n | 17.54% | 10.50% | 14.00% | 7.46% |
| **07** | BD-1 | 253 | 20 | 32 | y | 0.8 | n | y | y | n | -69.11% | -48.00% | -12.59% | -1.02% |
| **08** | BD-2 | 21 | 25 | 36 | n | - | y | y | n | n | -32.46% | -55.37% | 18.16% | 8.89% |
| **09** | BD-1 | 17 | 16 | 21 | y | 1.01 | n | y | n | n | -2.87% | 4.25% | -1.47% | 23.46% |
| **10** | BD-1 | 19 | 18 | 26 | n | - | n | y | y | n | 8.32% | 17.18% | 17.67% | 24.32% |

**Table S1. Demographic and SICI values of participants within the BDep group.**

*Yellow highlighted are values defined as outliers, using the 1.5* IQR method.*

*IQR = interquartile range, AUC=area under the curve, LMFP=local mean field power, GMFP=global mean field power, SICI=short latency intracortical inhibition, DLPFC=dorsolateral prefrontal cortex, BD=bipolar disorder, SIGH-ADS=Structured Interview Huide for Hamilton Depression rating scale – with Atypical Depression Supplement, HDRS=Hamilton Depression Rating Scale-17, AD = antidepressant medication, AP = antipsychotic medication, AE = anti-epileptic medication, Benzo = benzodiazepine.*
