## Supplementary Table 2 for "Left-sided Inhibition Deficit and right-sided Hyperexcitability in Treatment Resistant Bipolar Depression: A TMS-EEG study"

**Table S2. Spearman correlation tests between clinical variables and LMFP-AUC/GMFP-AUC after spTMS, ppTMS and the SICI (as ratio).**

|  | **Variable** | **Clinical variable** | **rho** | **p-value** |
| --- | --- | --- | --- | --- |
| **Left DLPFC** | | | | |
| **SICI** | **LMFP** | **HDRS** | -0.17 | 0.65 |
|  |  | **SIGH-ADS** | -0.31 | 0.39 |
|  |  | **Index bipolar** | 0.38 | 0.28 |
|  |  | **Index depression** | **-0.628** | **0.052** |
|  | **GMFP** | **HDRS** | -0.45 | 0.19 |
|  |  | **SIGH-ADS** | -0.53 | 0.11 |
|  |  | **Index bipolar** | 0.47 | 0.17 |
|  |  | **Index depression** | -0.51 | 0.13 |
| **spTMS** | **LMFP** | **HDRS** | -0.03 | 0.95 |
|  |  | **SIGH-ADS** | -0.12 | 0.74 |
|  |  | **Index bipolar** | 0.33 | 0.35 |
|  |  | **Index depression** | 0.012 | 0.97 |
|  | **GMFP** | **HDRS** | -0.12 | 0.74 |
|  |  | **SIGH-ADS** | -0.29 | 0.42 |
|  |  | **Index bipolar** | 0.50 | 0.14 |
|  |  | **Index depression** | -0.32 | 0.37 |
| **ppTMS** | **LMFP** | **HDRS** | -0.018 | 0.96 |
|  |  | **SIGH-ADS** | -0.018 | 0.96 |
|  |  | **Index bipolar** | 0.23 | 0.52 |
|  |  | **Index depression** | 0.24 | 0.51 |
|  | **GMFP** | **HDRS** | 0.27 | 0.45 |
|  |  | **SIGH-ADS** | 0.11 | 0.76 |
|  |  | **Index bipolar** | 0.47 | 0.17 |
|  |  | **Index depression** | -0.037 | 0.92 |
| **Right DLPFC** | | | | |
| **SICI** | **LMFP** | **HDRS** | 0.018 | 0.95 |
|  |  | **SIGH-ADS** | 0.049 | 0.89 |
|  |  | **Index bipolar** | 0.42 | 0.23 |
|  |  | **Index depression** | -0.21 | 0.55 |
|  | **GMFP** | **HDRS** | -0.19 | 0.60 |
|  |  | **SIGH-ADS** | -0.21 | 0.55 |
|  |  | **Index bipolar** | 0.28 | 0.43 |
|  |  | **Index depression** | -0.40 | 0.26 |
| **spTMS** | **LMFP** | **HDRS** | 0.45 | 0.19 |
|  |  | **SIGH-ADS** | 0.37 | 0.29 |
|  |  | **Index bipolar** | 0.26 | 0.47 |
|  |  | **Index depression** | -0.38 | 0.27 |
|  | **GMFP** | **HDRS** | **0.85** | **0.002** |
|  |  | **SIGH-ADS** | **0.66** | **0.036** |
|  |  | **Index bipolar** | -0.19 | 0.59 |
|  |  | **Index depression** | -0.53 | 0.11 |
| **ppTMS** | **LMFP** | **HDRS** | 0.44 | 0.21 |
|  |  | **SIGH-ADS** | 0.34 | 0.34 |
|  |  | **Index bipolar** | 0.085 | 0.82 |
|  |  | **Index depression** | -0.40 | 0.26 |
|  | **GMFP** | **HDRS** | **0.80** | **0.005** |
|  |  | **SIGH-ADS** | **0.59** | **0.075** |
|  |  | **Index bipolar** | -0.40 | 0.25 |
|  |  | **Index depression** | -0.15 | 0.69 |

*# = trend level, * = p≤0.05.*

*spTMS=single pulse Transcranial Magnetic Stimulation ppTMS=paired pulse Transcranial Magnetic Stimulation, AUC=area under the curve, LMFP=local mean field power, GMFP=global mean field power, DLPFC=dorsolateral prefrontal cortex, BD=bipolar disorder, HC = healthy control, Md=median. Statistical significance was reached when p≤0.05*
