## Supplementary Table 3 for "Left-sided Inhibition Deficit and right-sided Hyperexcitability in Treatment Resistant Bipolar Depression: A TMS-EEG study"

**Table S3. statistical analysis of SICI within group, between stimulation sides (left vs. right DLPFC).** Results are shown in median (IQR).

|  | **BD** | | | **HC** | | |
| --- | --- | --- | --- | --- | --- | --- |
|  | **Left-DLPFC** | **Right-DLPFC** | **Statistics** | **Left-DLPFC** | **Right-DLPFC** | **Statistics** |
| **Local SICI** | **3.7% (12.5%)** | **17.3%**  **(16.1%)** | **V=9**  **p=0.06^#^** | 9.0% (20.3%) | 0.75%  (27.2%) | V=123  p=0.11 |
| **Global SICI** | **1.5%**  **(11.7%)** | **16.2%**  **(14.6%)** | **V=1**  **p=0.004*** | 8.8% (20.6%) | -1.0%  (21.8%) | V=114  p=0.23 |

*# = trend level, * = p≤0.05*

*AUC=area under the curve, LMFP=local mean field power, GMFP=global mean field power, SICI=short latency intracortical inhibition, DLPFC=dorsolateral prefrontal cortex, BD=bipolar disorder group, HC = healthy control group. Statistical significance was reached when p≤0.05.*
