## Supplementary Table 4 for "Left-sided Inhibition Deficit and right-sided Hyperexcitability in Treatment Resistant Bipolar Depression: A TMS-EEG study"

**Table S4. Wilcoxon tests between individual TEPs of HC and BD, after spTMS and ppTMS, at the left and right DLPFC.** Results are shown in median (IQR).

|  | | **HC** | **BD** |  |
| --- | --- | --- | --- | --- |
| **Left DLPFC** | **Peak** | **Amp (uV)** | **Amp (uV)** | **Statistics** |
| **spTMS** | P30 | 0.042 (3.17) | 0.13 (1.95) | W=108, p=0.95 |
|  | N40 | -1.58 (2.42) | -1.27 (2.23) | W=119, p=0.73 |
|  | P60 | 1.68 (2.34) | 1.85 (3.75) | W=104, p=0.82 |
|  | N100 | -3.98 (4.76) | -4.04 (2.42) | W=100, p=0.98 |
|  | P180 | 7.60 (2.55) | 7.09 (3.70) | W=115, p=0.85 |
| **ppTMS** | P30 | 0.34 (1.89) | 0.41 (2.13) | W=119, p=0.73 |
|  | N40 | -0.02 (2.79) | 0.64 (2.23) | W=99, p=0.67 |
|  | P60 | 1.84 (2.56) | 2.46 (2.75) | W=93, p=0.50 |
|  | N100 | -4.60 (2.98) | -3.28 (2.25) | W=103, p=0.79 |
|  | P180 | 7.62 (4.39) | 6.84 (5.62) | W=107, p=0.92 |
| **Right DLPFC** | | | | |
| **spTMS** | P30 | 1.41 (1.88) | 1.42 (2.16) | W=100, p=0.65 |
|  | N40 | -0.52 (4.25) | -1.20 (1.74) | W=104, p=0.52 |
|  | P60 | 1.86 (3.70) | 2.65 (3.74) | W=74, p=0.46 |
|  | **N100** | **-3.34 (1.74)** | **-5.23 (6.42)** | **W=134, p=0.037** |
|  | P180 | 6.15 (3.96) | 6.51 (2.94) | W=81, p=0.68 |
| **ppTMS** | P30 | 0.86 (2.26) | 0.83 (2.17) | W=91, p=0.98 |
|  | N40 | 0.26 (1.71) | -0.26 (1.06) | W=108, p=0.40 |
|  | **P60** | **2.09 (2.28)** | **0.12 (4.09)** | **W=133, p=0.042** |
|  | N100 | -3.59 (1.81) | -4.65 (5.69) | W=99, p=0.68 |
|  | P180 | 5.68 (4.16) | 6.00 (3.23) | W=91, p=0.98 |

*# = trend level, * = p≤0.05.*

*spTMS=single pulse Transcranial Magnetic Stimulation, ppTMS=paired pulse Transcranial Magnetic Stimulation DLPFC=dorsolateral prefrontal cortex, BD=bipolar disorder, HC = healthy control, Md=median. Statistical significance was reached when p≤0.05.*
